# Association Between Use of Renin-Angiotensin System Inhibitors and Cardiovascular Outcomes Following Transcatheter Aortic Valve Implantation: A Systematic Review and Meta-Analysis

**DOI:** 10.64898/2026.05.07.26352703

**Authors:** Daniel J. Nyberg, Christopher S. Dodgson, John M. Aalen, Christian H. Eek, Bjørn Bendz, Lars Aaberge, Peder L. Myhre, Kristoffer E. Russell, Øyvind H. Lie

## Abstract

**Background:** Aortic stenosis (AS) carries a high mortality risk if left untreated. Transcatheter aortic valve implantation (TAVI) has emerged as a primary treatment modality for many patients with severe AS. Observational data suggest that renin-angiotensin system (RAS) inhibitor use following TAVI are associated with lower risk, but with divergent reported effects and limited statistical power for specific cardiovascular outcome. This study aimed to assess the association between RAS inhibitor use and clinical outcomes after TAVI.

**Methods:** A systematic literature search was conducted in EMBASE and PubMed. Eligible studies included those reporting on RAS inhibitor use in TAVI populations. The primary outcome was all-cause mortality. Secondary outcomes included cardiovascular mortality, myocardial infarction (MI), cerebrovascular events, and heart failure (HF) hospitalization.

**Results:** Nine observational studies including 36,015 patients were included. RAS inhibitor use was associated with lower odds of all-cause mortality (OR 0.74, 95% CI 0.70– 0.78), cardiovascular mortality (OR 0.62, 95% CI 0.55–0.72), cerebrovascular events (OR 0.59, 95% CI 0.47–0.74), and HF hospitalization (OR 0.84, 95% CI 0.77–0.92). No clear association was observed for MI (OR 0.95, 95% CI 0.59–1.53).

**Conclusions:** RAS inhibitor use was associated with favorable clinical outcomes following TAVI. However, these findings are based on observational data, which are subject to residual confounding. Randomized controlled trials are needed to clarify the clinical utility of RAS inhibitors in this setting.

## Introduction

Aortic stenosis (AS) is the most common valvular heart disease in older adults, affecting approximately one-third of individuals over the age of 65.(1) Once symptomatic, AS is associated with high mortality if left untreated. Treatment options include surgical aortic valve replacement (SAVR) and transcatheter aortic valve implantation (TAVI), with the latter increasingly used across a broader age and risk spectrum. Earlier data demonstrates an elevated mortality in patients receiving TAVI compared to the general population. However, this mortality excess appears to be attenuated in contemporary practice, possibly due to procedural advancements and evolving patient selection. (2,3) Optimizing long-term medical therapy after TAVI remains a priority.

Current guideline-directed therapy post-TAVI is limited to antithrombotic management.(4) Patients with indications for oral anticoagulation, such as atrial fibrillation, typically receive a non-vitamin K antagonist oral anticoagulant (NOAC), while others are treated with a single antiplatelet agent. One randomized clinical trial reported a benefit of dapagliflozin after TAVI.(5) Evidence supporting additional pharmacotherapy to improve long-term outcomes is lacking.

Renin-angiotensin system (RAS) inhibitors, including angiotensin-converting enzyme inhibitors (ACEi) and angiotensin II receptor blockers (ARB), have demonstrated efficacy in patients with heart failure, hypertension, and chronic kidney disease.(6–8) Many of these comorbidities are prevalent, yet often underdiagnosed and untreated in patients with severe AS. The widespread availability, low cost and favorable safety profile further support interest in evaluating a potential role following TAVI. Observational data suggests potential benefits of RAS inhibitors in this setting. However, the evidence remains inconclusive due to divergent effect estimates across studies, and limited statistical power for analyzing specific cardiovascular outcomes. Furthermore, concerns regarding tolerability, especially in elderly patients with comorbidities and polypharmacy, highlight the need for a comprehensive evaluation.

This systematic review and meta-analysis aimed to investigate the association between RAS inhibitor use and cardiovascular outcomes in patients with AS undergoing TAVI.

## Methods

### Search Strategy and Study Selection

A systematic literature review was performed in EMBASE and PubMed between January 8 and 12, 2024, to identify studies reporting on RAS inhibitor use in patients undergoing TAVI. The search combined terms related to “renin-angiotensin system,” “ACE inhibitors,” “angiotensin receptor blockers,” and “TAVI/TAVR,” applying Boolean logic and filters for human studies. The goal was to retrieve the most comprehensive set of relevant literature.

Inclusion criteria followed the PICO framework: Population—patients undergoing TAVI for AS; Intervention—treatment with RAS inhibitors; Comparator—no RAS inhibitor treatment; Outcome—all-cause mortality (primary). Secondary outcomes included cardiovascular mortality, MI, HF hospitalization, and cerebrovascular events.

### Outcomes

While the primary objective of this meta-analysis was to evaluate associations between RAS inhibitor use and cardiovascular outcomes post-TAVI, all-cause mortality was designated as the primary endpoint due to the substantial competing risks in this population. Secondary outcomes encompassed cardiovascular mortality, myocardial infarction, cerebrovascular events, and heart failure hospitalization.

### Data Extraction and Statistical Analysis

Event rates in the control groups were estimated using reported cumulative proportions. Meta-analyses were performed using Peto’s method (common effects model), and results were expressed as weighted mean average odds ratios (OR) with 95% confidence intervals (CI). Heterogeneity was assessed using I^2^ statistics. A funnel plot was constructed to evaluate the possible impact of publication bias. All analyses were conducted using RStudio (version 4.3.1) with the “meta” package.

## Results

### Study Characteristics

The search yielded 148 records. After removing duplicates and screening titles, abstracts, and full texts, nine observational studies were included, encompassing 36,015 patients (9–17)(figure 1). No randomized controlled trials were identified at the time of the search. The included studies varied in design: most were retrospective cohorts; one was prospective, and one used propensity score matching.

**Figure 1.**
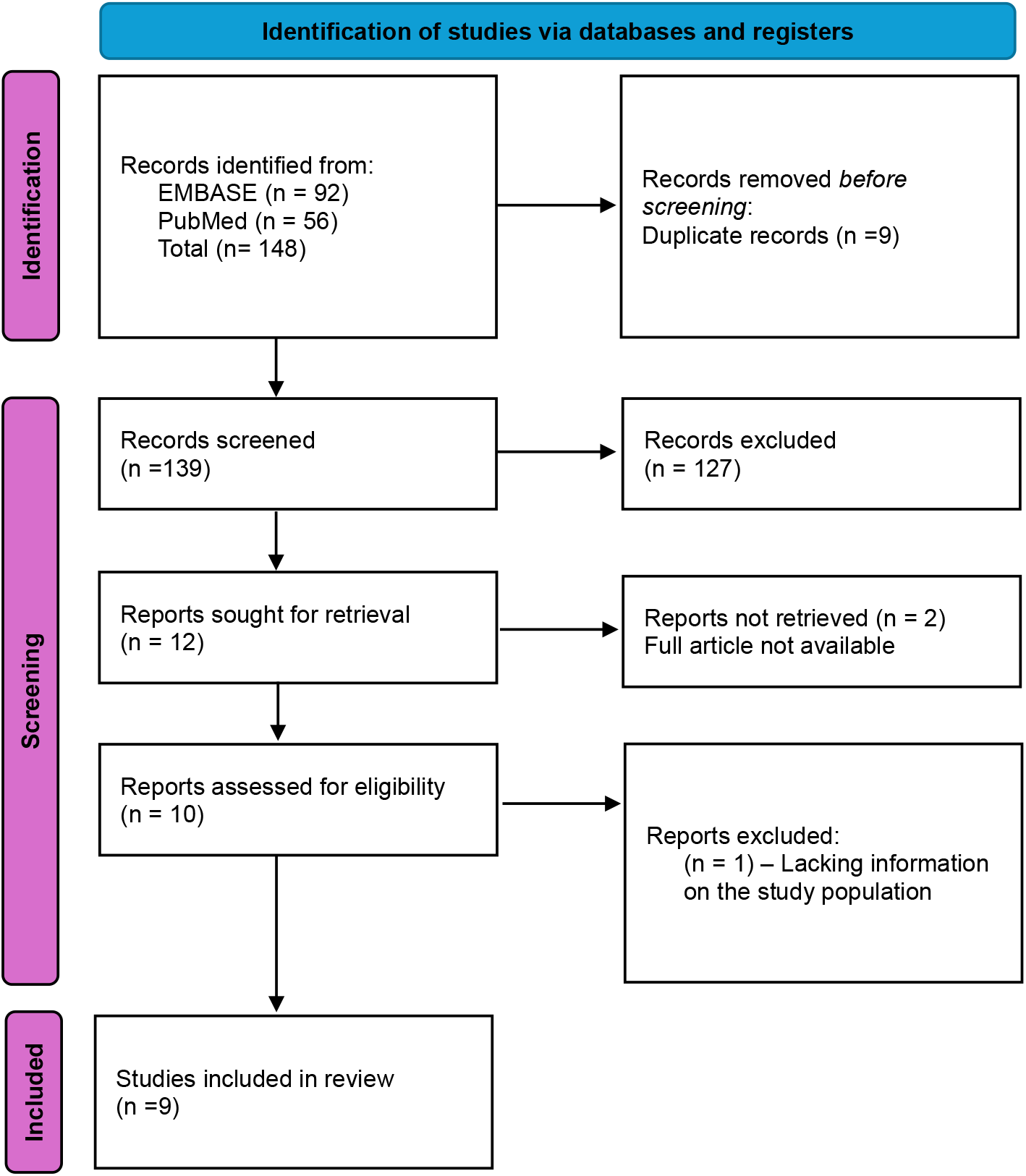
Consort diagram of selection of eligible published analyses.

In total 17 565 (49%) patients used RAS inhibitors after TAVI, consisting primarily of ACEi or ARB. Mineralcorticoid receptor antagonists (MRA) were included as RAS inhibitors in two studies in a small minority of patients (6% and 3,3%). (12,14) Patient characteristics were generally similar across studies. Mean age ranged from 79.1 to 84.5 years (table 1), 74.8% to 97.5% had hypertension and 37.3% to 75.1% had chronic kidney disease. Patients in the RAS inhibitor groups typically had a higher prevalence of hypertension, diabetes mellitus and coronary artery disease. One study made use of propensity score matching in an effort to balance baseline characteristics.(16) Patient characteristics are summarized in table 1.

**Table 1.** Reported patient characteristics (RASi vs no-RASi)

| Study | n= RASi vs no RASi |  | Age RASi vs no-RASi |  | Sex (% female) RASi vs no-RASi |  | Hypertension RASi vs no-RASi |  | Diabetes mellitus RASi vs no-RASi |  | Coronary artery disease RASi vs no-RASi |  | Chronic kidney disease, eGFR≤60 mL/min/1.73 m<sup>2</sup> RASi vs no-RASi |  |
| --- | --- | --- | --- | --- | --- | --- | --- | --- | --- | --- | --- | --- | --- | --- |
| Fischer-Rasokat U et.al | 2227 | 635 | 82.0 | 82.0 | 52.2 % | 52.9 % | 93.4 % | 82.0 % | 32.9 % | 32.6 % | 60.9 % | 53.4 % | n.r* |  |
| Klinkhammer B et.al | 71 | 98 | 77.8 | 80.1 | 43.7 % | 38.8 % | 95.8 % | 76.5 % | 50.7 % | 31.6 % | 74.6 % | 76.5 % | 46.5 % | 57.1 % |
| Ledwoch J et.al | 225 | 98 | 80.6 | 79.8 | 45.9 % | 44.0 % | 94.2 % | 81.1 % | 25.8 % | 18.9 % | 75.6 % | 60.4 % | n.r* |  |
| Kaewkes D et.al | 349 | 415 | 81.4 | 82.9 | 43.0 % | 39.5 % | 93.7 % | 86.5 % | 34.1 % | 24.1 % | 42.7 % | 39.0 % | 75.1 % | 73.0 % |
| Chen S et.al | 1736 | 2243 | 81.7 | 82.9 | 39.2 % | 41.6 % | 96.3 % | 89.7 % | 40.6 % | 32.5 % | 81.0 % | 76.0 % | n.r* |  |
| Rodriguez-Gabella T et.al | 1622 | 1163 | 80.8 | 80.7 | 54.9 % | 53.1 % | 85.6 % | 74.8 % | 36.4 % | 31.7 % | 39.1 % | 32.9 % | 37.3 % | 42.5 % |
| Ochiai T et.al | 371 | 189 | 84.2 | 84.8 | 68.2 % | 65.6 % | 83.8 % | 61.4 % | 27.8 % | 24.3 % | 41.5 % | 33.9 % | n.r* |  |
| Inohara T et.al | 7948 | 7948 | 82.4 | 82.4 | 47.7 % | 48.4 % | 93.6 % | 93.1 % | 38.7 % | 38.7 % | n.r** |  | 46.7 % | 46.2 % |
| Cubeddu R et.al | 3172 | 5840 | 79.3 | 80.4 | 48.1 % | 45.6 % | 97.5 % | 92.8 % | 50.6 % | 41.8 % | 86.8 % | 86.4 % | n.r* |  |

\*Investigators did not report chronic kidney disease as eGFR≤60 mL/min/1.73 m<sup>2</sup>
\*\*Investigators did not report coronary artery disease as one collective disease, but as prevalence of individual conditions such as myocardial infarction, triple vessel disease and left main disease

### Meta-Analysis Findings

RAS inhibitor use was associated with a significant 26% lower odds of all-cause mortality (OR 0.74, 95% CI 0.70–0.78, Figure 2). For secondary outcomes, RAS inhibitor use was linked to reduced odds of cardiovascular mortality (OR 0.62, 95% CI 0.55–0.72, Figure 3), cerebrovascular events (OR 0.59, 95% CI 0.47–0.74, Figure 4) and HF hospitalization (OR 0.84, 95% CI 0.77–0.92, Figure 4). There was no clear association with altered odds of MI (OR 0.95, 95% CI 0.59–1.53, Figure 4).

**Figure 2.**
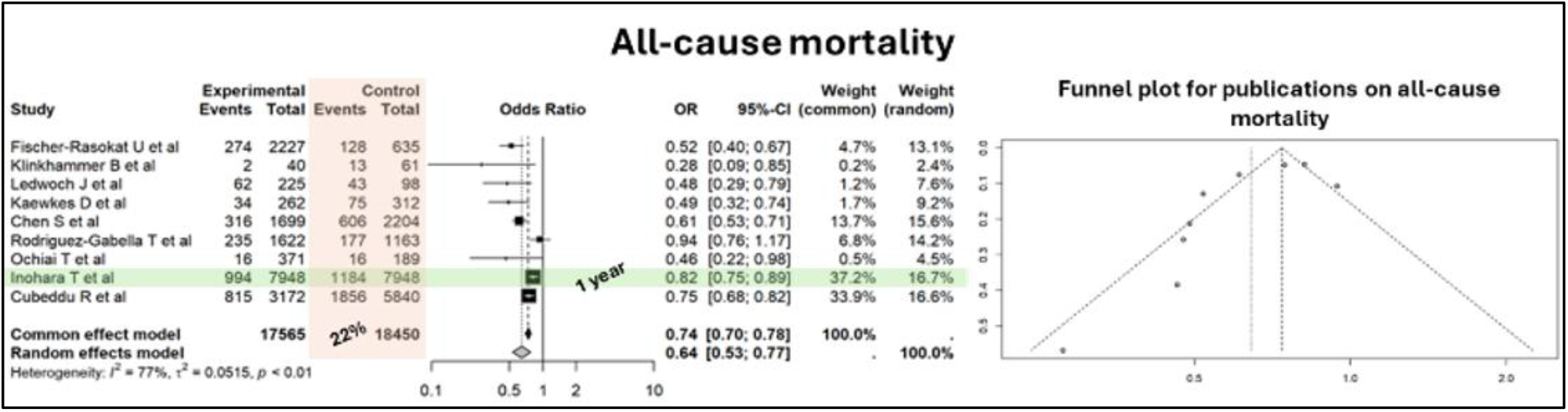
All-cause mortality in users and non-users of RAS-inhibitors after TAVI Figure legend: Forest plot (left panel) of tabular metanalysis of all-cause mortality after TAVI in patients with and without RAS-inhibitors and the corresponding funnel plot for publication bias. Green color highlights reports with shorter follow-up duration. Orange color highlights the mean reported prevalence of all-cause mortality in the control groups. Funnel plot (right panel) describes the distribution of the reported effect estimates (X-axis) to their standard error (Y-axis). (CI = confidence interval, OR = odds ratio, RAS-inhibitors = renin angiotensin system inhibitors, TAVI = transcatheter aortic valve implantation)

**Figure 3.**
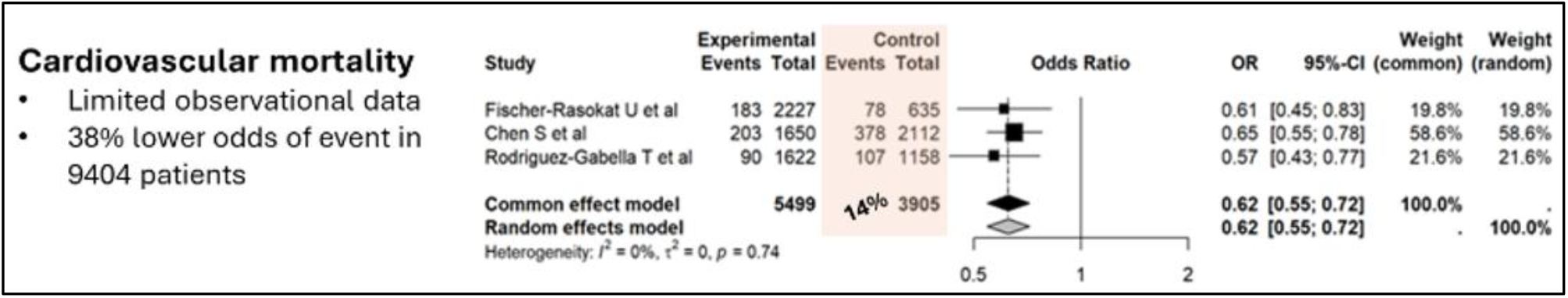
Cardiovascular mortality in users and non-users of RAS-inhibitors after TAVI Figure legend: Forest plot of tabular metanalysis of cardiovascular mortality after TAVI in patients with and without RAS-inhibitors. Orange color highlights the mean reported prevalence of cardiovascular mortality in the control groups. (CI = confidence interval, OR = odds ratio, RAS-inhibitors = renin angiotensin system inhibitors, TAVI = transcatheter aortic valve implantation)

**Figure 4.**
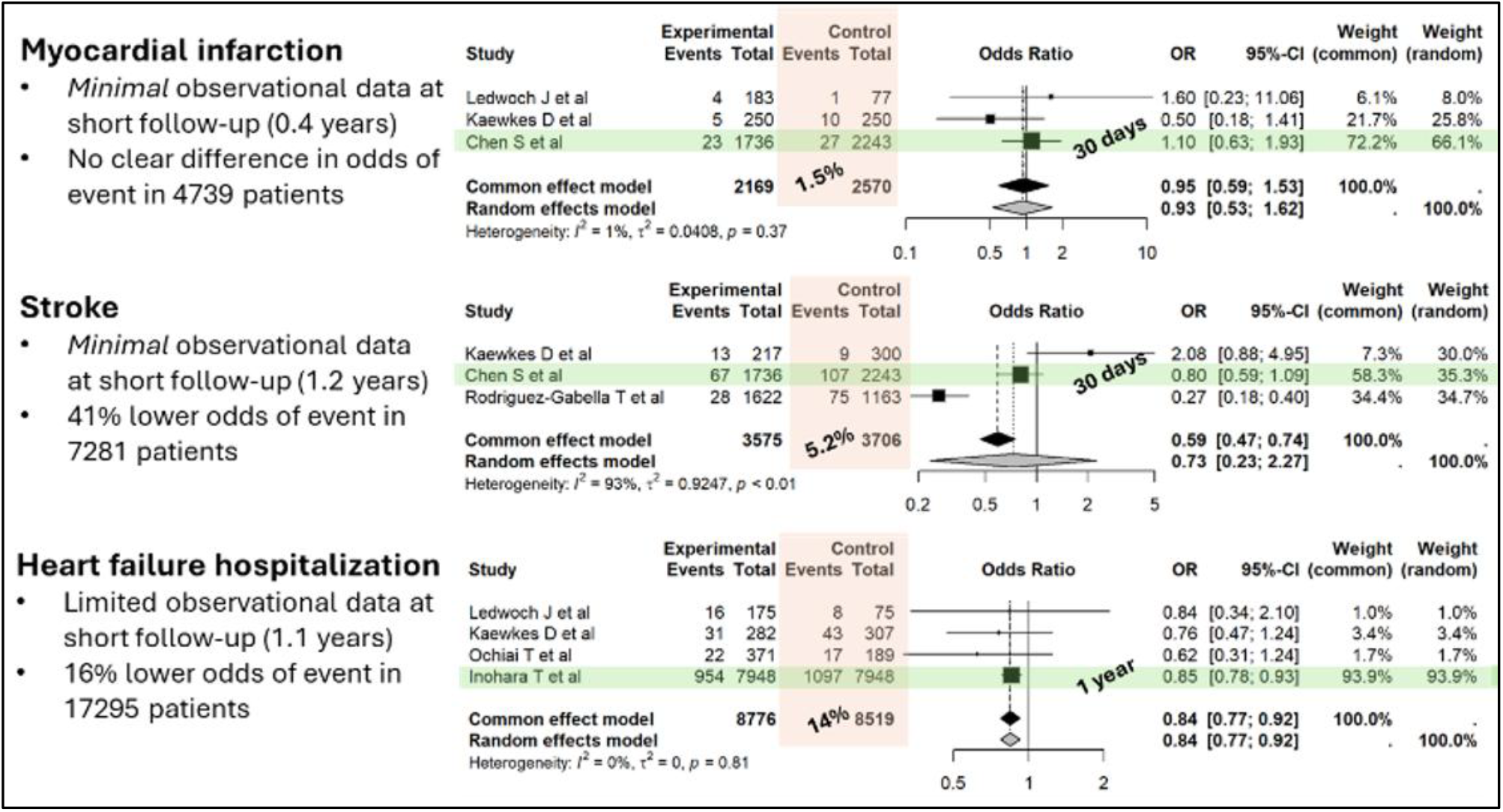
Secondary outcome measures in users and non-users of RAS-inhibitors after TAVI Figure legend: Forest plot of tabular metanalyses of secondary outcomes after TAVI in patients with and without RAS-inhibitors. Green color highlights reports with shorter follow-up duration. Orange color highlights the prevalence of secondary outcome event rates in the control groups. (CI = confidence interval, OR = odds ratio, RAS-inhibitors = renin angiotensin system inhibitors, TAVI = transcatheter aortic valve implantation)

Heterogeneity was moderate to high for some outcomes (I^2^ = 77% for all-cause mortality; I^2^ = 93% for cerebrovascular events). A funnel plot for reported odds ratios for all-cause mortality indicated asymmetry, suggesting potential publication bias favoring positive findings (Figure 2).

## Discussion

This meta-analysis suggests an association between RAS inhibitor use and improved outcomes following TAVI, including reduced all-cause and cardiovascular mortality, as well as fewer cerebrovascular events and HF hospitalizations. No clear association was found for MI.

Several pharmacological mechanisms may explain these findings. Improved blood pressure control and reverse cardiac remodeling through attenuation of neurohormonal activation are potential contributors. RAS blockers are cornerstone therapy in patients with HF, with studies suggesting benefits not only in HF with reduced ejection fraction (HFrEF), but in patients with LVEF up to 50%.(18) In patients with severe AS, HF is common, but often unrecognized. Signs and symptoms are frequently attributed to the valvular disease, rather than HF itself. Moreover, many clinicians are reluctant to initiate RAS blockers in the setting of severe AS due to concerns about hypotension from vasodilation. This undertreatment may persist even after correcting the AS with TAVI, potentially withholding important therapy with prognostic significance.

Despite methodological limitations, the results of this systematic review and meta-analysis demonstrate an interesting phenomenon that relates to the direction of the most obvious biases. The findings are notable given that RAS inhibitors are typically prescribed to patients with higher comorbidity burdens. The most common indications are hypertension, heart failure and renal disease. These conditions are all associated with worse clinical outcomes in patients with AS. Intuitively, the RAS inhibitor group could therefore be expected to have worse clinical outcomes than the patients not treated with RAS inhibitors. The RAS inhibitor groups in the included studies had consistently higher prevalence of hypertension, diabetes mellitus and coronary artery disease than the groups without RAS inhibitors. Another possible bias to the disadvantage of the intervention is the follow-up time. The total mean follow-up time in the studies was generally short, and ranged from 0,4-2,6 years between the different outcomes. Considering the possible mechanisms for RAS inhibitors, one could speculate that benefits would occur during longer term follow-up after TAVI.

Despite the potential confounders that work against beneficial effects of RAS inhibitors, our meta-analysis clearly shows that patients treated with this class of drug have lower event rates. However, selection bias may also explain the findings. Patients in the RAS inhibitor group received their medication based on clinical judgement. Clinicians may have chosen to prescribe these drugs to patients they perceived to be more robust. The decision to withhold RAS blocker therapy in patients who were believed not to tolerate the drugs could therefore introduce confounding by indication. The underlying frailty confers worse prognosis, not the treatment or lack there-of. Additionally, patients who tolerated and continued RAS inhibitors after initiation may represent a healthier subset, introducing survivorship bias. However, patients in both groups have initially been considered candidates for TAVI, which may suggest a reasonable overall health status.

The funnel plot for publications on all-cause mortality suggests a bias towards publication of smaller studies with signs of benefit. Considering possible missing studies showing neutral results or negative effects for RAS inhibitors, the effect sizes in the present meta-analysis are likely imprecise.

The RASTAVI trial is the only RCT testing the effect of RAS inhibitors after TAVI. In this trial, 186 patients who had undergone TAVI and had LVEF >40% were randomized to ramipril or standard of care. After one year follow-up the primary composite endpoint of cardiac mortality, heart failure readmission, and stroke was similar between the groups (10.6% vs 12.0%, p=0.78). However, the trial was underpowered, particularly to detect individual outcome differences.(19) Although the present meta-analysis provide important observational data supporting the use of RAS inhibitors after TAVI, methodological limitations, residual confounding and potential biases make the results unsuitable to guide clinical decision making. A large, adequately powered clinical outcomes trial is needed.

### Limitations

The primary limitation of this study is its reliance on observational data which are susceptible to residual confounding and biases. Heterogeneity across studies and varying follow-up durations further complicate interpretation. Dosage and timing of RAS inhibitor therapy were inconsistently reported, and pre-TAVI RAS inhibitor status was not uniformly addressed.

## Conclusions

This systematic review and meta-analysis found that RAS inhibitor use following TAVI is associated with reduced odds of all-cause mortality, cardiovascular mortality, cerebrovascular events, and HF hospitalizations. The observational nature of the data limits the ability to infer causality. These findings support the need for a randomized controlled trial to evaluate the clinical utility of RAS inhibitors in this patient population.

## Data Availability

All data are available upon request

